# 30-day mortality and morbidity in COVID-19 versus influenza: A population- based study

**DOI:** 10.1101/2020.07.25.20162156

**Authors:** Vardan Nersesjan, Moshgan Amiri, Hanne Christensen, Michael E. Benros, Daniel Kondziella

**Author notes:** contributed equally as first authors (Vardan Nersesjan, Moshgan Amiri). contributed equally as senior authors (Michael E. Benros, Daniel Kondziella). **Corresponding author:** Daniel Kondziella, MD, MSc, PhD, FEBN, Department of Neurology (section 2083), Rigshospitalet, Copenhagen University Hospital, DK-2100 Copenhagen.

## Abstract

**Background:** As of July 2020, COVID-19 has caused 500,000 deaths worldwide. However, large-scale studies of COVID-19 mortality and new-onset comorbidity compared to influenza and individuals tested negative for COVID-19 are lacking. We aimed to investigate COVID-19 30-day mortality and new-onset comorbidity compared to individuals with negative COVID-19 test results and individuals tested for influenza.

**Methods and findings:** This population-based cohort study utilized electronic health records covering roughly half (n=2,647,229) of Denmark’s population, with nationwide linkage of microbiology test results and death records. All individuals ≥18 years tested for COVID-19 and individuals tested for influenza were followed from November 1, 2017 to June 30, 2020. The main outcome was 30-day mortality after a test for either COVID-19 or influenza. Secondary outcomes were major comorbidity diagnoses 30-days after the test for either COVID-19 or influenza. In total, 224,639 individuals were tested for COVID-19. Among inpatients positive for COVID-19, 356 of 1657 (21%) died within 30 days, which was a 3.0 to 3.1-fold increased 30-day mortality rate, when compared to influenza and COVID-19-negative inpatients (all p<0.001). For outpatients, 128 of 6,263 (2%) COVID-19-positive patients died within 30 days, which was a 5.5 to 6.9-fold increased mortality rate compared to influenza and COVID-19-negative patients, respectively (all p<0.001). Compared to hospitalized patients with influenza, new-onset ischemic stroke, diabetes and nephropathy occurred more frequently in inpatients with COVID-19 (all p<0.05).

**Conclusions:** In this population-based study comparing COVID-19 with influenza, COVID-19 was associated with increased rates of major systemic and vascular comorbidity and substantially higher mortality, which is likely even higher than the stated 3.0 to 5.5-fold increase owing to more extensive testing for COVID-19.

## 1. INTRODUCTION

COVID-19 has led to a worldwide healthcare crisis with >10,000,000 confirmed infected people, resulting in >500,000 deaths as of July 2020.^1,2^ Governmental initiatives including lockdowns and social distancing are aiming to restrict the spread of the virus. However, critical voices^3^ have argued the socioeconomic consequences may be unjustified given that little is known about how the pandemic compares with annual influenza epidemics in terms of mortality and morbidity. According to the WHO, seasonal influenza may result in 290,000-650,000 deaths worldwide annually,^4,5^ but substantially higher mortality rates for COVID-19 might have even more adverse impact on global health without strict preventive measures.

Of further concern, COVID-19 might not only be a respiratory disease but a multi-organ disorder because of the wide expression of the angiotensin-converting enzyme-2 receptor to which SARS-Cov-2 binds,^6^ leading among others to thromboembolic complications,^7^ severe inflammatory responses^8^ and possibly diabetes.^9^ Neurological and psychiatric complications will likely constitute a major health burden as well.^10,11^ But how COVID-19 morbidity compares to influenza morbidity is equally poorly understood.

Here, for the first time, we utilized population-based electronic health records (EHR) from Denmark linked with nationwide databases on test results for infections and death records, to investigate mortality in people with COVID-19 compared to people with influenza and to people tested negative for COVID-19. For secondary outcomes we estimated COVID-19-associated new-onset comorbidity, including cardiovascular, neurological and psychiatric events, compared to influenza and individuals tested COVID-19-negative. Analyses were stratified according to age and in- and outpatient status. We hypothesized that COVID-19 would be associated with higher mortality and increased rates of novel comorbidities compared to influenza A/B.

## 2. METHODS

This retrospective Danish study was based on EHR covering two well-defined administrative regions: Capital Region (i.e. Greater Copenhagen and Bornholm) and Region Zealand, comprising roughly 50% of the Danish population. Denmark has an almost exclusively public health care sector based on catchment areas.

### Registers and study population

The EHR system of the Capital and Zealand Regions, EPIC (Verona, Wisconsin, USA), consists of data from all hospital contacts in these regions. From implementation in 2016 to June 30, 2020, 2,647,229 individuals were registered. Diagnoses are defined according to ICD-10.^12^ Registration of death in the EHR is synchronized with the Danish national population registry, updated every 24h. Accuracy of test results for influenza and SARS-CoV-2 virus is ensured by synchronization of EPIC with the nationwide Danish Microbiology Database.^13^ All individuals ≥18 years tested for COVID-19 between March 1-June 1, 2020, and all individuals tested for influenza A/B between November 1, 2017-June 1, 2020, were followed for mortality and new-onset comorbidities 30-days after the test until June 30, 2020.

### Assessment of COVID-19 and influenza test results

#### COVID-19

All individuals tested for COVID-19 during March 1-June 1, 2020 with laboratory tests CORONAVIRUS 2019-NCOV and/or CORONAVIRUS SARS-COV-2 RNA via nasal, pharyngeal and/or tracheal samples with reverse-transcriptase-polymerase-chain reaction (RT-PCR) assays were included. These specific tests cover all performed COVID-19 tests in the catchment areas and are available from the Danish Microbiology Database^13^.

#### Influenza A/B

We included all individuals tested for influenza A/B during November 1, 2017 to March 1, 2020, using 9 different RT-PCR laboratory tests (***eTable 1***), covering all available influenza tests based on nasal, pharyngeal and/or tracheal samples.

**Table 1.** Demographics and prior comorbidities among individuals tested for COVID-19 or influenza as in- or outpatient.

|  | Inpatients |  |  | Outpatients |  |  |
| --- | --- | --- | --- | --- | --- | --- |
|  | COVID-19 positive<br>(N = 1,657) | COVID-negative<br>(N = 31,483) | Influenza-positive<br>(N = 7,200) | COVID-19-positive<br>(N = 6,263) | COVID-negative<br>(N = 158,400) | Influenza-positive<br>(N = 7,204) |
| **Age – years, no. (%)** |  |  |  |  |  |  |
| Mean, years | 65 | 60 | 66 | 47 | 48 | 49 |
| 18 - 39 | 182 (11.0) | 7,456 (23.7)<sup>‡</sup> | 931 (12.9) | 2,310 (36.9) | 54,659 (34.5)<sup>‡</sup> | 2,390 (33.2)<sup>‡</sup> |
| 40 - 59 | 427 (25.8) | 6,790 (21.6)<sup>‡</sup> | 1,413 (19.6)<sup>‡</sup> | 2,509 (40.1) | 62,701 (39.6) | 2,637 (36.6)<sup>‡</sup> |
| 60 - 80 | 636 (38.4) | 11,261 (35.8)<sup>‡</sup> | 2,876 (39.9) | 1,043 (16.7) | 33,166 (20.9)<sup>‡</sup> | 1,762 (24.4)<sup>‡</sup> |
| > 80 | 412 (24.9) | 5,976 (19.0)<sup>‡</sup> | 1,980 (27.5) | 401 (6.4) | 7,874 (5.0)<sup>‡</sup> | 415 (5.8) |
| **Sex (%)** |  |  |  |  |  |  |
| Women | 737 (44.5) | 17,099 (54.3)<sup>‡</sup> | 3,844 (53.4)<sup>‡</sup> | 3,931 (62.8) | 99,364 (62.7) | 4,272 (59.3)<sup>‡</sup> |
| **Prior medical diagnoses - no. (%) #** |  |  |  |  |  |  |
| Neurological, any | 275 (16.6) | 4,998 (15.9) | 1,025 (14.2) | 505 (8.1) | 14,074 (8.9)<sup>‡</sup> | 474 (6.6)<sup>‡</sup> |
| Cerebrovascular, any | 141 (8.5) | 2,819 (9.0) | 485 (6.7) | 145 (2.3) | 3,698 (2.3) | 125 (1.7)<sup>‡</sup> |
| Ischemic stroke incl. TIA | 78 (4.7) | 1,739 (5.5) | 165 (2.3)<sup>‡</sup> | 85 (1.4) | 2,302 (1.5) | 130 (1.8)<sup>‡</sup> |
| Psychiatric, any | 202 (12.2) | 6,581 (20.9)<sup>‡</sup> | 821 (11.4) | 383 (6.1) | 12,264 (7.7)<sup>‡</sup> | 305 (4.2)<sup>‡</sup> |
| Ischemic heart disease | 147 (8.9) | 2,631 (8.4) | 547 (7.6) | 90 (1.4) | 3,719 (2.3)<sup>‡</sup> | 155 (2.2)<sup>‡</sup> |
| Heart failure | 100 (6.0) | 1,861 (5.9) | 422 (5.9) | 45 (0.7) | 1,581 (1.0)<sup>‡</sup> | 68 (0.9) |
| Diabetes | 199 (12.0) | 3,291 (10.5)<sup>‡</sup> | 827 (11.5) | 209 (3.3) | 4,903 (3.1) | 255 (3.5) |
| Chronic lower respiratory disease | 182 (11.9) | 3,943 (12.5) | 1,362 (18.9)<sup>‡</sup> | 223 (3.6) | 7,934 (5.0)<sup>‡</sup> | 366 (5.1)<sup>‡</sup> |
| Obesity | 46 (2.8) | 1,296 (4.1)<sup>‡</sup> | 167 (2.3) | 132 (2.1) | 3,929 (2.5) | 141 (2.1) |
| COVID-19: positive COVID-19 PCR test between March 1-June 1, 2020; influenza: positive influenza A/B PCR test between November 1, 2017 to March 1, 2020; COVID-neg: negative COVID-19 PCR test between March 1-June 1, 2020. Each patient was followed for a total of 30 days from positive test until end of follow-up or death. * Indicates statistically significant difference (p<0.05) compared to COVID-19 populations. # Established medical diagnoses, registered in the medical files, prior to testing for COVID-19 or influenza. TIA, transitory ischemic attack. |  |  |  |  |  |  |

### Outcome measures

#### Primary outcome measures

30-day mortality in the COVID-19-positive population, compared to 30-day mortality of the control population with COVID-19-negative tests and the influenza-positive and –negative populations.

#### Secondary outcome measures

New-onset (i.e. 30 days after COVID-19 or influenza test) comorbidity diagnoses, including neurological, psychiatric and cardiovascular disease, pulmonary embolism, venous thrombosis, renal failure, diabetes and rheumatoid arthritis, in all populations. ICD-10 codes are listed in **eTable 1**.

### Data collection, statistical analysis and ethics

Anonymized retrospective aggregate-level data on sex, age, prior comorbidities and population mortality 30 days after test results were extracted for individuals ≥18 years for each population, using the EPIC Slicer-Dicer function. For search strategies see **eTable 1**. As individuals could be tested multiple times, individuals were only included in the COVID-19-negative, respectively, influenza-negative populations, when all their tests had been negative. Individuals tested for influenza during March 1-June 1, 2020 (i.e. FLU-19) were included for sensitivity analysis (*see below*). To avoid overlap, we removed COVID-19-positive individuals from the FLU-19 population.

Main analysis was the relative risk (RR) of mortality rates 30 days after a test, in the overall populations and stratified according to in- and outpatient status and age. Secondary analysis was RR of cumulative 30 days post-test incidence of new-onset comorbidities, after exclusion of individuals who already had the investigated comorbidity before the test. We compared COVID-19 positive with COVID-19-negative and influenza-positive individuals. To validate mortality data, absolute mortality rates extracted from electronic health records (EPIC) were compared with official Danish statistics numbers (**eTable 2**). Sensitivity analysis was conducted by comparing individuals ≥18 years with a positive or negative influenza test from the same time period as the COVID-19 population, i.e. March 1-June 1, 2020 (FLU-19), in order to investigate the possible influence of the COVID-19 pandemic, including lockdown and social distancing measures, on mortality rates in individuals tested for influenza. Chi-squared statistics were used to calculate odds ratio (OR), RR and 95% confidence intervals (CI) using SPSS (version 25; IBM, Armonk, NY, USA). Two-sided p≤0.05 was considered significant.

**Table 2.** Relative risk of 30-day mortality after a COVID-19 or influenza test among in- or outpatients.

|  |  |  |  |  |  |  |
| --- | --- | --- | --- | --- | --- | --- |
|  | *Inpatients* |  |  | *Outpatients* |  |  |
|  | Total (N) | Death (N) | RR (95% CI) | Total (N) | Death (N) | RR (95% CI) |
| **COVID-19-positive vs. COVID-negative** |  |  |  |  |  |  |
| **Overall** | 1,657 vs 31,483 | 356 vs 2,185 | **3.10** (2.80-3.42)<sup>†</sup> | 6,263 vs 158,400 | 128 vs 469 | **6.90** (5.69-8.38)<sup>†</sup> |
| **18-39 years** | 182 vs 7,456 | ≤5 vs 15 | N/A | 2,310 vs 54,659 | ≤5 vs ≤5 | N/A |
| **40-59 years** | 427 vs 6,790 | 16 vs 158 | 1.61 (0.97-2.67) | 2,509 vs 62,701 | ≤5 vs 20 | N/A |
| **60-80 years** | 636 vs 11,261 | 150 vs 1,004 | **2.65** (2.27-3.08)<sup>†</sup> | 1,043 vs 33,166 | 33 vs 161 | **6.52** (4.50-9.43)<sup>†</sup> |
| **> 80 years** | 412 vs 5,976 | 190 vs 1,008 | **2.73** (2.42-3.08)<sup>†</sup> | 401 vs 7,874 | 92 vs 288 | **6.27** (5.07-7.76)<sup>†</sup> |
| **COVID-19 positive vs. FLU-positive** |  |  |  |  |  |  |
| **Overall** | 1,657 vs 7,200 | 356 vs 516 | **3.00** (2.65-3.39)<sup>†</sup> | 6,263 vs 7,204 | 128 vs 27 | **5.45** (3.61-8.25)<sup>†</sup> |
| **18-39 years** | 182 vs 931 | ≤5 vs ≤5 | N/A | 2,310 vs 2,390 | ≤5 vs ≤5 | N/A |
| **40-59 years** | 427 vs 1,413 | 16 vs 26 | **2.04** (1.10-3.76)<sup>‡</sup> | 2,509 vs 2,638 | ≤5 vs ≤5 | N/A |
| **60-80 years** | 636 vs 2,876 | 150 vs 214 | **3.17** (2.62-3.83)<sup>†</sup> | 1,043 vs 1,761 | 33 vs ≤5 | N/A |
| **> 80 years** | 412 vs 1,980 | 190 vs 271 | **3.37** (2.89-3.92)<sup>†</sup> | 401 vs 415 | 92 vs 20 | **4.76** (2.30-7.57)<sup>†</sup> |
| **COVID-19 positive vs. FLU-negative** |  |  |  |  |  |  |
| **Overall** | 1,657 vs 26,366 | 356 vs 2,873 | **1.97** (1.79-2.18)<sup>†</sup> | 6,263 vs 19,314 | 128 vs 129 | **3.06** (2.40-3.90)<sup>†</sup> |
| **18-39 years** | 182 vs 3,867 | ≤5 vs 40 | N/A | 2,310 vs 6,664 | ≤5 vs ≤5 | N/A |
| **40-59 years** | 427 vs 4,757 | 16 vs 224 | 0.79 (0.48-1.31) | 2,509 vs 6,454 | ≤5 vs 6 | N/A |
| **60-80 years** | 636 vs 10,939 | 150 vs 1,307 | **1.97** (1.70-2.30)<sup>†</sup> | 1,043 vs 4,963 | 33 vs 51 | **3.08** (2.00-4.75)<sup>†</sup> |
| **> 80 years** | 412 vs 6,803 | 190 vs 1,302 | **2.41** (2.15-2.70)<sup>†</sup> | 401 vs 1,233 | 92 vs 71 | **3.98** (2.98-5.32)<sup>†</sup> |
| COVID-19, positive COVID-19 test between March 1-June 1, 2020; COVID-neg, negative COVID-19 test between March 1-June 1, 2020; FLU positive, positive influenza A/B test between November 1, 2017-March 1, 2020; FLU negative, negative influenza A/B test between November 1, 2017-March 1, 2020. Each included patient was followed for a total of 30 days from positive test until end of follow-up or death. N/A, not applicable due to low number of patients (N≤5). † p < 0.001; ‡ p < 0.05 |  |  |  |  |  |  |

The Ethics Committee of the Capital Region of Denmark waives approval for register-based studies on aggregated anonymized data (Section 14.2 of the Committee Act. 2; http://www.nvk.dk/english). Use of anonymized aggregate-level data was approved by the Danish Data Protection Agency. Results from ≤5 patients were displayed as “≤5” to ensure data privacy.

## 3. RESULTS

A total of 224,639 individuals of any age were tested for SARS-CoV-2 between March 1-June 1, 2020; positive results were found in 7,920 individuals ≥18 years (i.e. our case population). A negative COVID-19 test occured in 189,883 individuals ≥18 years. Between November 1, 2017-March 1, 2020, we identified 79,414 individuals, who were tested for influenza A/B. Positive results were found in 14,404 individuals aged ≥18 years. Negative influenza A/B tests were identified in 45,680 individuals ≥18 years (**eFigure 1**). Demographics are displayed in **Table 1** and **eTables 3-5**. The proportion of inpatients at the time of COVID-19 or influenza tests was lower in the COVID-19-positive (20.9%) and the COVID-19-negative (16.6%) populations compared to influenza-positive (50%) and influenza-negative (57.7%) populations. We therefore analyzed mortality and comorbidities both in the overall populations and stratified according to in- and outpatient status and age.

**Figure 1.**
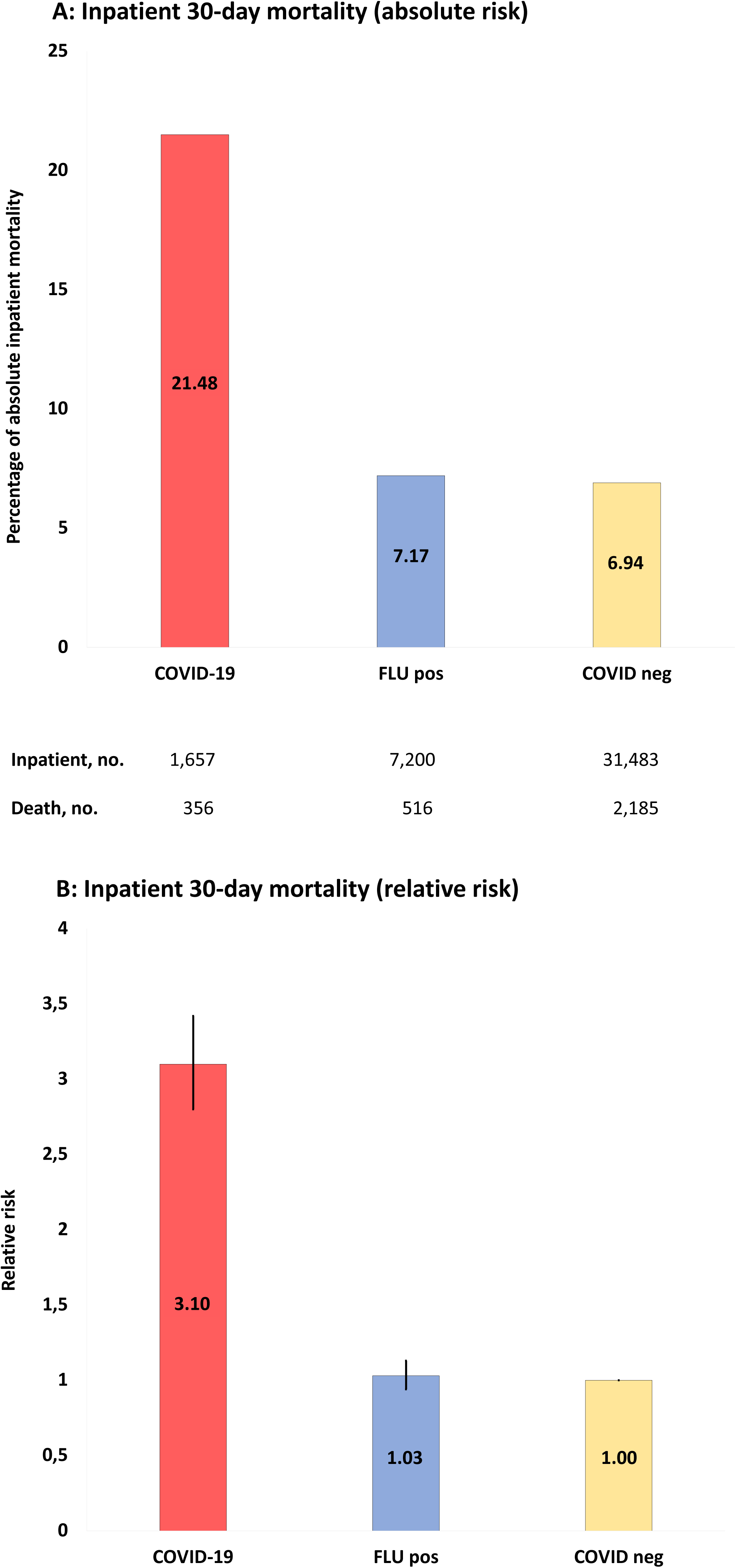
**A:** Absolute risk with 95% CI of COVID-19 inpatient 30-day mortality, when compared to populations of the study: COVID-19, positive COVID-19 test between March 1-June 1, 2020; FLU pos, positive influenza A/B test between November 1, 2017-March 1, 2020; COVID-neg, negative COVID-19 test between March 1-June 1, 2020. **B:** RR with 95% CI of inpatient mortality of study populations compared to COVID-19 negative population as reference.

### Primary outcome: Mortality

#### Overall mortality rates

Overall 30-day mortality in COVID-19-positive individuals was 484 of 7,920 (6.1%), whereas 30-day mortality for COVID-19-negative individuals was 2,654 of 189,883 (1.4%), corresponding to an increased mortality by RR 4.37 (95% CI=3.98-4.80).

#### Mortality rates of inpatients tested for COVID-19 and/or influenza

30-day mortality for hospitalized COVID-19 patients ≥18 years was 356 of 1,657 (21.5%), which was higher than in COVID-19-negative individuals (30-day mortality 2,185/31,483; 6.9%; p<0.001) (**Figure 1, Table 2** and **eTable 6-9**). The corresponding numbers for individuals tested positive for influenza were 516/7,200 (7.2%) and for influenza-negative individuals 2873/26,366 (11%). Mortality for COVID-19-positive inpatients was increased by RR 3.10 (95% CI=2.80-3.42) compared to COVID-19-negative patients, and by RR 3.00 (95% CI=2.65-3.39) and RR 1.97 (95% CI=1.79-2.18) compared to influenza-positive, respectively, influenza-negative inpatients (all p<0.001).

When mortality rates were stratified according to age, 30-day mortality rates for hospitalized COVID-19 patients were 16/427 (3.7%, age 40-59 years), 150/636 (23.6%, 60-80 years) and 190/412 (46%, >80 years). The corresponding numbers for COVID-19-negative individuals were 158/6,790 (2.3%), 1,004/11,261 (8.9%) and 1,008/5,976 (16.9%) and for influenza-positive individuals 26/1,413 (1.8%), 214/2,876 (7.4%) and 271/1,980 (13.7%), respectively. Mortality for COVID-19-positive inpatients was significantly increased with age 60-80 years (RR=2.65; 95% CI=2.27-3.08) and >80 years (RR=3.17; 95% CI=2.62-3.83), when compared to COVID-19-negative individuals (RR 2.73; 95% CI=2.42-3.08) and influenza-positive individuals (RR 3.37 (95% CI=2.89-3.92). See **Table 2** and **eTable 6** for a full outline of inpatient mortality rates stratified according to age groups.

#### Mortality rates of outpatients tested for COVID-19 and/or influenza

Regarding outpatients, positive COVID-19 tests were associated with 128 deaths in 6,263 people (2% 30-day mortality) and negative COVID-19 tests with 469 deaths in 158,400 people (0.3%), whereas the corresponding numbers for influenza-tested people were 27/7,204 (0.4%; positive test) and 129/19,314 (0.7%; negative test). Mortality rates for COVID-19-positive outpatients were increased by RR 6.90 (95% CI=5.69-8.38) compared to COVID-19-negative outpatients, by RR 5.45 (95% CI=3.61-8.25) compared to influenza-positive outpatients, and by RR 3.06 (95% CI=2.40-3.90) compared to influenza-negative outpatients. **Figure 2** and **eTable 6-9** show details.

**Figure 2.**
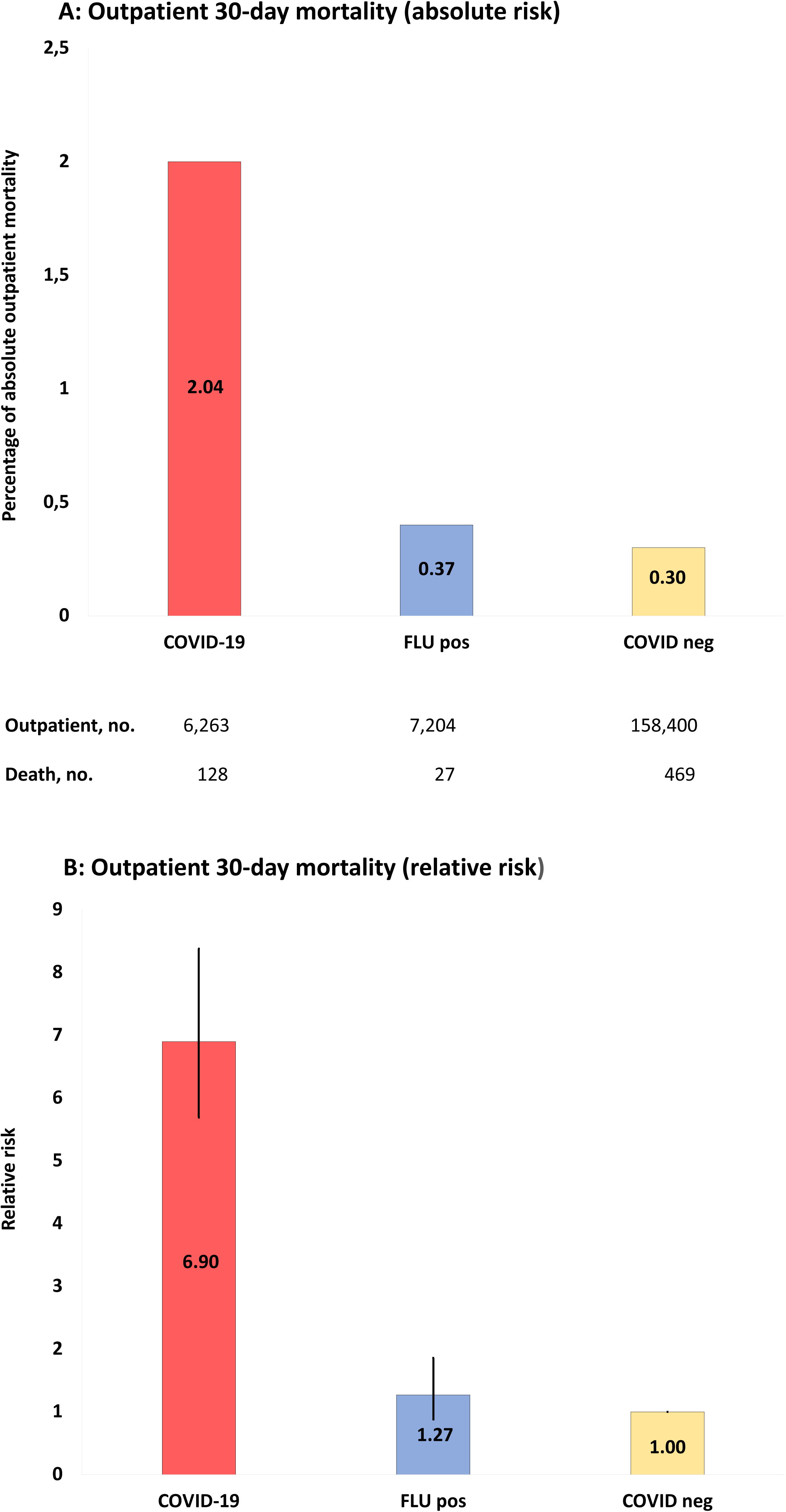
**A:** Absolute risk with 95% CI of COVID-19 outpatient 30-day mortality, when compared to populations of the study: COVID-19, positive COVID-19 test between March 1-June 1, 2020; FLU pos, positive influenza A/B test between November 1, 2017-March 1, 2020; COVID-neg, negative COVID-19 test between March 1-June 1, 2020. **B:** RR with 95% CI of outpatient mortality of study populations compared to COVID-19 negative population as reference.

The 30-day mortality rates for outpatients with COVID-19 were 20/62,701 (0.03%), 33/1,043 (3.2%) and 92/401 (22.9%) for age groups 40-59, 60-80 and >80 years, respectively. The corresponding numbers for COVID-19 negative individuals were 161/33,166 (0.5%) and 288/7,874 (3.7%) in the age groups 60-80 and >80 years, and for influenza-positive individuals ≤5/1,761 and 20/415 (4.8%), respectively. The case numbers were too low in the remaining age groups for statistics. See **Table 2** and **eTable 6** for full details of outpatient mortality rates stratified according to age.

### Secondary outcomes: New-onset comorbidities

**Figure 3** and **eTables 10-11** display data regarding novel diagnoses after COVID-19 and influenza tests.

**Figure 3.**
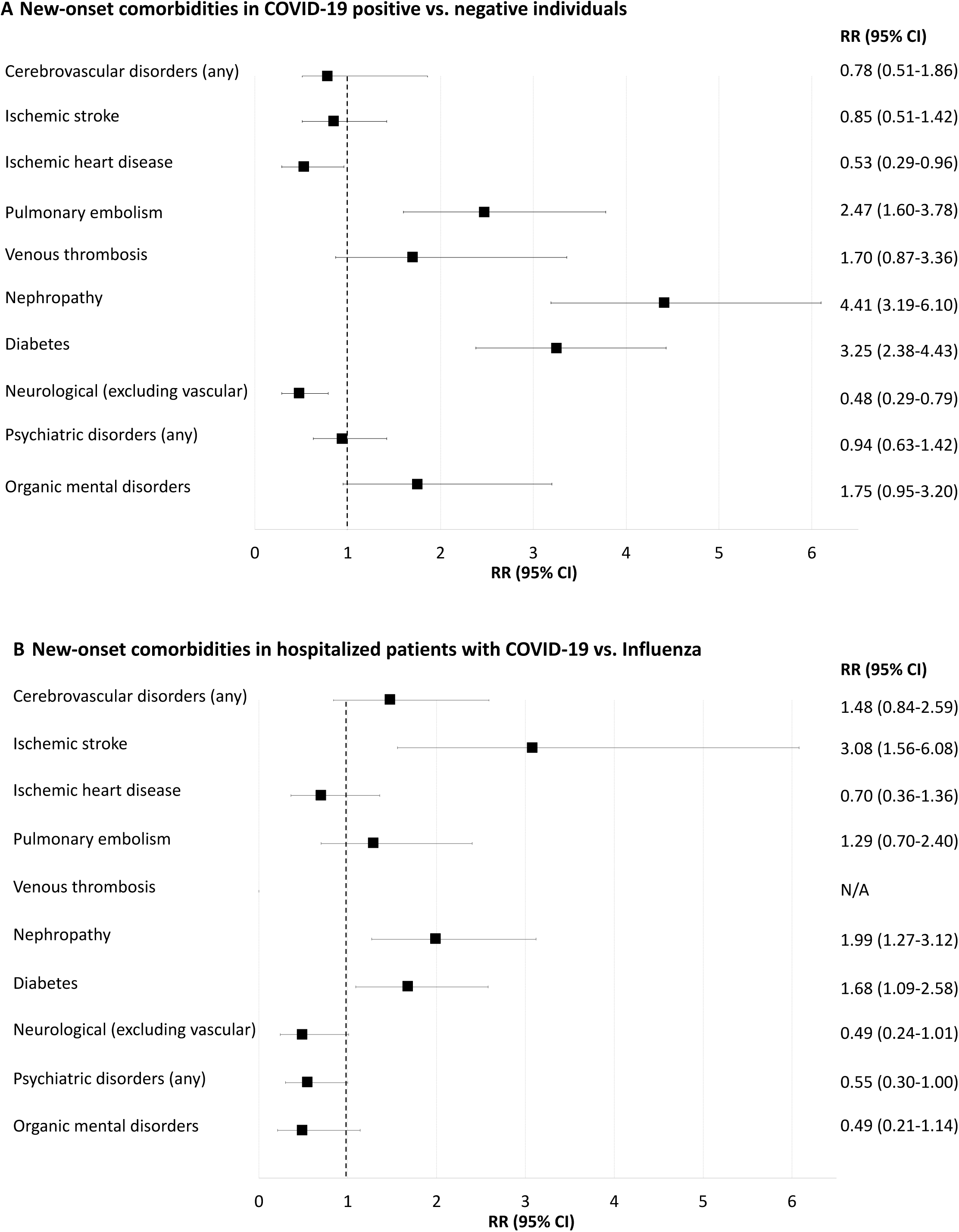
Forest-plot showing relative risk of new-onset comorbidities 30 days after positive or negative COVID-19 or influenza A/B tests (i.e. prior comorbidities excluded). **A**: COVID-19 positive compared to COVID-19 negative individuals. **B**: Inpatients with positive COVID-19 compared to inpatients with positive influenza. N/A, not applicable because of too few cases for meaningful statistics (≤ 5 individuals). New-onset delirium, neurotic and anxiety disorders, affective disorders, schizophrenia and delusional disorders and rheumatoid arthritis yielded too few cases for meaningful statistics (data shown in **eTable 10-11**).

#### New-onset comorbidities among COVID-19-positive and COVID-19-negative individuals

Pulmonary embolism 30 days after testing was more frequent in COVID-19-positive compared to COVID-19-negative individuals (RR 2.47 (95% CI=1.60-3.78)), **eTable 10**. Diabetes and renal failure were also more frequent in COVID-19-positive compared to negative individuals (0.6% vs. 0.2% and 0.6% vs. 0.1%, respectively; both p<0.001). Neurological disorders (excluding vascular disorders) and ischemic heart disease were less frequent in COVID-19-positive than in COVID-19-negative people (0.2% vs. 0.5% and 0.1% vs. 0.3%, respectively; both p< 0.05). Rates of new-onset cerebrovascular disorders, venous thrombosis and psychiatric disorders were not significantly different between the two populations.

#### New-onset comorbidities in inpatients tested positive for COVID-19 versus influenza-positive individuals

Incident ischemic stroke 30 days after a test was more frequent in COVID-19-positive inpatients compared to those with influenza, RR 3.10 (95% CI=1.56-6.08), **eTable 11**. New-onset diabetes and nephropathy were more frequent in COVID-19 positive compared to influenza-positive inpatients (1.9% vs. 1.2% and 1.8% vs. 0.9% respectively; both p< 0.05). Rates of new-onset pulmonary embolism, neurological disorders and psychiatric disorders were not statistically different.

#### New-onset comorbidities in outpatients tested positive for COVID-19 versus influenza-positive individuals

Incidence diagnoses 30 days after positive tests in outpatients yielded either too low numbers for meaningful statistics or were not statistically different (**eTable 11)**.

### Sensitivity analysis

The COVID-19-positive population was compared to a population of influenza-tested individuals from the same time period, March 1-June 1, 2020, i.e. outside the influenza peak season (FLU-19). In total, 12,502 people were tested for influenza A/B (56% inpatients; 566 positive and 8,318 negative). Inpatient mortality in FLU-19-positive and -negative populations was 26/317 (8.2%), respectively, 578/5058 (11.4%). Inpatient mortality was significantly increased in COVID-19 compared to FLU-19-positive and -negative individuals (RR 2.62 (95% CI=1.79-3.83), respectively, RR 1.88 (95% CI=1.67-2.12); both p<0.001).

## 4. DISCUSSION

To our knowledge, this is the first population-based study comparing mortality rates and new-onset comorbidities of COVID-19 patients with those of influenza patients and COVID-19-negative controls. 30-day mortality was 3.0 to 6.9-fold higher in the positive COVID-19 population compared to influenza and COVID-19-negative control populations. The largest difference in mortality between COVID-19 and influenza was observed in outpatients. Equally important, new-onset ischemic stroke, renal failure and diabetes occurred at increased rates in COVID-19-positive inpatients compared to influenza patients.

Previous studies have reported widely varying overall COVID-19 mortality rates, e.g. 1.4% among 1099 cases in Wuhan, China,^14^ and 7.2% among 22,512 in Italy.^15^ In our study, the overall 30-day COVID-19 mortality was 6.1%, which is well in line with previous data from COVID-19 patients from Denmark.^16^ Importantly, mortality rates are very different among in- and outpatients. In the COVID-19-positive inpatient population 30-day mortality was 21%, corresponding well again with a mortality of 28% in 191 inpatients reported by Zhou et al.,^17^ respectively, a median 14-day mortality of 26% in 140 inpatients from Xie et al.^18^ These numbers are much higher than the 2% mortality in outpatients (i.e. individuals from the general public not requiring hospitalization) in the present study, indicating that, not surprisingly, inpatients with COVID-19 are doing worse than outpatients.

Compared to influenza and COVID-19-negative populations, COVID-19 30-day mortality was increased 3.0 to 3.10-fold for inpatients and 5.5 to 6.9-fold for outpatients. This is somewhat in contrast with an estimated 20-fold mean increase of COVID-19 mortality compared to influenza, based on indirect estimated numbers from the general public in the US.^19^ This discrepancy could be explained by the higher proportion of sick individuals in our influenza tested populations. If testing for influenza A/B in Denmark had been equally widespread as for COVID-19, the excess COVID-19 mortality gap would likely have been even larger. As expected, mortality in COVID-19-positive patients was lowest in the young and highest in the elderly.

Thromboembolic complications in COVID-19 are assumed to be frequent.^20^ New-onset ischemic stroke was indeed more frequent in COVID-19 than in influenza inpatients. Increased rates of ischemic stroke in COVID-19 compared to influenza were also found in another study based on retrospective medical charts review from 2 academic centers in New York.^21^ Given that signs and symptoms of stroke – especially minor stroke – may be obscured by systemic illness as well as sedation and ventilation, the true risk may even be higher than the 3-to 7-fold increase reported here and in the cited work.^21^ We also found that the risk of new-onset diabetes was 3-fold elevated in COVID-19-positive individuals compared to negative controls and 2-fold elevated compared to influenza-positive patients. These results substantiate concerns of diabetogenic effects of COVID-19,^22^ including the possibility of ketoacidosis.^23^ Similarly, nephropathy was frequent in our COVID-19 population, and renal failure may lead to more complications and higher in-hospital mortality.^24^ Ischemic heart disease appeared equally prevalent in inpatients with COVID-19 and those with influenza. Finally, pulmonary embolism occurred more often in our COVID-19 positive population compared to negative controls (albeit not compared to influenza populations).

All these comorbidities, alone or in combination, may put patients with COVID-19 at risk for multiorgan failure. This, together with hypoxemia owing to pulmonary changes, including diffuse alveolar damage with fibrin membranes, thickened alveolar walls, lymphocytic infiltration,^25^ and pulmonary thrombosis,^25^ complicated by cardiac arrhythmias, hypotensive shock,^26^ and possibly brainstem dysfunction,^27^ is being proposed as the final pathway to death in COVID-19.^28^ Many of these mechanisms are unlikely to be specific enough to be reliably captured by diagnostic coding in EHR-based studies such as ours. Large prospective multicenter registries and autopsy studies comparing COVID-19 patients with COVID-19-negative controls and influenza victims are required to dissect the exact contribution of each of these factors.

Concerns for neurological and psychiatric complications in COVID-19 are increasingly being raised.^11^ Yet, most (albeit not all^10^) reports have revealed a predominance of relatively unspecific symptoms such as altered mental state in highly selected groups,^11,29,30^ while we report on EHR-registered diagnoses. Our results show decreased or similar frequencies of new-onset neurological and psychiatric diagnoses in COVID-19 individuals within 30 days of testing compared to influenza, which suggests either that these complications in COVID-19 are no more frequent than in influenza or that the nationwide lockdown in Denmark resulted in fewer contacts to the health care system by people with COVID-19 but relatively mild comorbid symptoms, including neurological and psychiatric ones. Indeed, observations from California, Italy and Denmark ^31–33^ indicate a lower incidence of hospitalization of patients with e.g. cardiac disease during the COVID-19 lockdown. Further, mild cognitive and emotional symptoms are not likely to be reported within 30 days.

Strengths of our study are related, among others, to the large population numbers and the catchment area-based approach. The extracted general mortality data during years 2018-2020 corresponded well to Danish statistics mortality data (**eTable 2**). Numbers of COVID-19 and influenza tests, test results, admissions, and mortality rates in this study were equally consistent with the official Danish numbers.^34^ Further, test results of SARS-CoV-2 and influenza swabs are synchronized with the Danish national microbiology database;^13^ and we validated our data extraction strategy by ensuring that two individual searches supervised by two independent Epic Slicer-Dicer experts yielded identical results. As to limitations, we were unable to adjust for confounding factors such as socioeconomic, lifestyle and ethnicity, owing to the use of aggregated EHR data which depend on correct coding by physicians. Selection bias might be considerable because individuals were tested in hospital settings (even as outpatients), and the testing strategy of COVID-19 in Denmark has been much more comprehensive compared to influenza.

### Conclusions

In this first population-based study comparing COVID-19 with influenza, COVID-19 was associated with substantially higher mortality, which is likely even higher than the stated 3.0-5.5-fold increase owing to more extensive testing for COVID-19, and we observed higher rates of new-onset ischemic stroke, diabetes and renal failure. Next, middle- and long-term follow-up data are required to investigate mortality trajectories in COVID-19 versus influenza populations, and molecular and genetic studies will have to elucidate the specific biological mechanisms behind COVID-19’s higher mortality and morbidity compared to influenza.

## Data Availability

Raw data will be made available upon reasonable request

## Acknowledgements

This work received funding from Lundbeckfonden (grant number R349-2020-658 and R268-2016-3925), RH Forskningspulje (R143-A6132-B3632), Region Hovedstadens Forskningsfond til Sundhedsforskning 2019 (A6597), Savværksejer Jeppe Juhl og Hustru Ovita Juhls Mindelegat (27062019), and Offerfonden (F-23101-04).The sponsor had no role in the acquisition of the data, interpretation of the results or the decision to publish the findings. ***Conflict of Interest Disclosures:*** The authors have no relevant financial disclosures.

## Supplementary Appendix

**eFigure 1.**
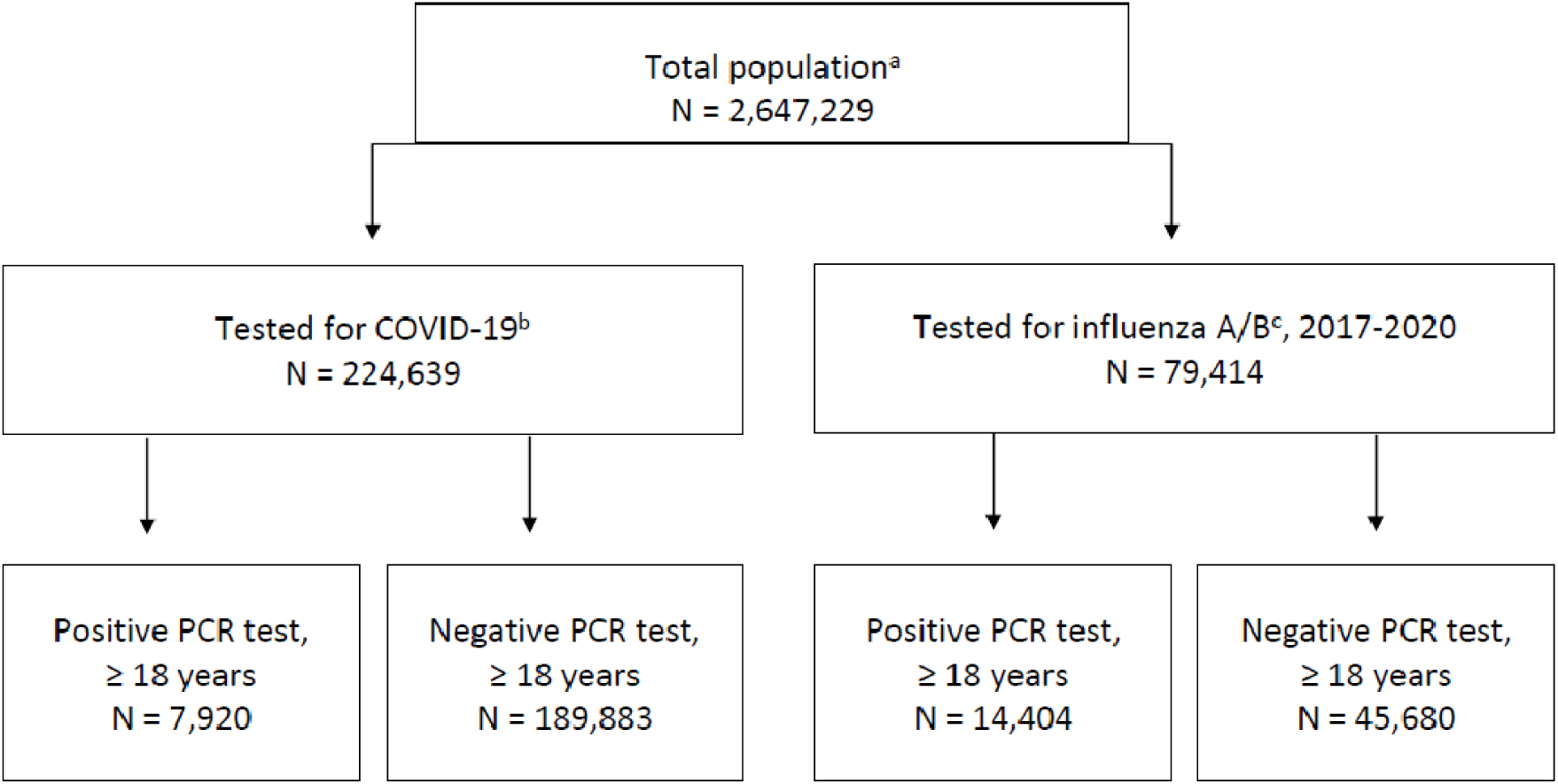
Flow chart of study populations. See Methods and Results for details. a total population in EHR registered per June 30, 2020; b tested between March 1-June 1, 2020; c tested between November 1, 2017-March 1, 2020.

**eTable 1.** Detailed search string utilized in EPIC SLICER DICER.

eTables 1-11
| eTable 1. Detailed search string utilized in EPIC SLICER DICER |  |  |
| --- | --- | --- |
| POPULATIONS | SEARCH METHOD | TIME PERIOD |
| COVID-19 |  |  |
| Positive | All patients ≥ 18 years **AND** (Laboratory findings of Positive SARS-COV-2 test **OR** CORONAVIRUS 2019-NCOV) | March 1 – June 1, 2020 |
| Negative | All patients ≥ 18 years **AND** (Laboratory findings of Negative SARS-COV-2 test **OR** CORONAVIRUS 2019-NCOV) **AND** (exclusion of positive SARS-COV-2 test **OR** CORONAVIRUS 2019-NCOV) | March 1 – June 1, 2020 |
| Influenza A/B |  |  |
| Positive, FLU | All patients ≥ 18 years **AND** (Positive laboratory findings of, INFLUENZA A, INFLUENZA B, INFLUENZA A RNA, INFLUENZA B RNA, INFLUENZA VIRUS B, INFLUENZA TYPE A, INFLUENZA TYPE B, INFLUENZA A RNA (POC) **OR** INFLUENZA B RNA (POC)) | November 1, 2017 – March 1, 2020 |
| Positive, FLU17 | All patients ≥ 18 years **AND** (Positive laboratory findings of, INFLUENZA A, INFLUENZA B, INFLUENZA A RNA, INFLUENZA B RNA, INFLUENZA VIRUS B, INFLUENZA TYPE A, INFLUENZA TYPE B, INFLUENZA A RNA (POC) **OR** INFLUENZA B RNA (POC)) | November 1, 2017 – March 1, 2018 |
| Positive, FLU18 | All patients ≥ 18 years **AND** (Positive laboratory findings of, INFLUENZA A, INFLUENZA B, INFLUENZA A RNA, INFLUENZA B RNA, INFLUENZA VIRUS B, INFLUENZA TYPE A, INFLUENZA TYPE B, INFLUENZA A RNA (POC) **OR** INFLUENZA B RNA (POC)) | November 1, 2018 – March 1, 2019 |
| Positive, FLU19 | All patients ≥ 18 years **AND** (Positive laboratory findings of, INFLUENZA A, INFLUENZA B, INFLUENZA A RNA, INFLUENZA B RNA, INFLUENZA VIRUS B, INFLUENZA TYPE A, INFLUENZA TYPE B, INFLUENZA A RNA (POC) **OR** INFLUENZA B RNA (POC)) **AND** (exclusion of Positive laboratory findings of SARS-COV-2 test **OR** CORONAVIRUS 2019-NCOV) | March 1 – June 1, 2020 |
| Negative, FLU | All patients ≥ 18 years **AND** (Negative laboratory findings of, INFLUENZA A, INFLUENZA B, INFLUENZA A RNA, INFLUENZA B RNA, INFLUENZA VIRUS B, INFLUENZA TYPE A, INFLUENZA TYPE B, INFLUENZA A RNA (POC) **OR** INFLUENZA B RNA (POC)) **AND** (exclusion of Positive laboratory findings of, INFLUENZA A, INFLUENZA B, INFLUENZA A RNA, INFLUENZA B RNA, INFLUENZA VIRUS B, INFLUENZA TYPE A, INFLUENZA TYPE B, INFLUENZA A RNA (POC) **OR** INFLUENZA B RNA (POC)) | November 1, 2017 – March 1, 2020 |
| Negative, FLU17 | All patients ≥ 18 years **AND** (Negative laboratory findings of, INFLUENZA A, INFLUENZA B, INFLUENZA A RNA, INFLUENZA B RNA, INFLUENZA VIRUS B, INFLUENZA TYPE A, INFLUENZA TYPE B, INFLUENZA A RNA (POC) **OR** INFLUENZA B RNA (POC)) **AND** (exclusion of Positive laboratory findings of, INFLUENZA A, INFLUENZA B, INFLUENZA A RNA, INFLUENZA B RNA, INFLUENZA VIRUS B, INFLUENZA TYPE A, INFLUENZA TYPE B, INFLUENZA A RNA (POC) **OR** INFLUENZA B RNA (POC)) | November 1, 2017 – March 1, 2018 |
| Negative, FLU18 | All patients ≥ 18 years **AND** (Negative laboratory findings of, INFLUENZA A, INFLUENZA B, INFLUENZA A RNA, INFLUENZA B RNA, INFLUENZA VIRUS B, INFLUENZA TYPE A, INFLUENZA TYPE B, INFLUENZA A RNA (POC) **OR** INFLUENZA B RNA (POC)) **AND** (exclusion of Positive laboratory findings of, INFLUENZA A, INFLUENZA B, INFLUENZA A RNA, INFLUENZA B RNA, INFLUENZA VIRUS B, INFLUENZA TYPE A, INFLUENZA TYPE B, INFLUENZA A RNA (POC) **OR** INFLUENZA B RNA (POC)) | November 1, 2018 – March 1, 2019 |
| Negative, FLU19 | All patients ≥ 18 years **AND** (Negative laboratory findings of, INFLUENZA A, INFLUENZA B, INFLUENZA A RNA, INFLUENZA B RNA, INFLUENZA VIRUS B, INFLUENZA TYPE A, INFLUENZA TYPE B, INFLUENZA A RNA (POC) **OR** INFLUENZA B RNA (POC)) **AND** (exclusion of Positive laboratory findings of, INFLUENZA A, INFLUENZA B, INFLUENZA A RNA, INFLUENZA B RNA, INFLUENZA VIRUS B, INFLUENZA TYPE A, INFLUENZA TYPE B, INFLUENZA A RNA (POC) **OR** INFLUENZA B RNA (POC)) **AND** (exclusion of Positive laboratory findings of SARS-COV-2 test **OR** CORONAVIRUS 2019-NCOV) | March 1 – June 1, 2020 |
| **DEMOGRAPHICS, mortality and prior/new-onset comorbidities** |  |  |
| SEX |  |  |
| Female / male | Use of slicing function for sex distribution of male or female on all POPULATIONS |  |
| Age |  |  |
| Mean | Use of measure function to calculate mean age on all POPULATIONS |  |
| Range | Use of slicing function for age range on all POPULATIONS |  |
| Admission status |  |  |
| Inpatient | POPULATION as defined **PLUS** overlapping criteria of hospitalization at the same time as laboratory test is performed |  |
| Outpatient | POPULATION as defined **AND** (exclusion of individuals with a corresponding laboratory test **PLUS** overlapping criteria of hospitalization at the same time as laboratory test is performed) |  |
| Mortality |  |  |
|  | POPULATION as defined **PLUS** sequential criteria of death up to 30 days after COVID-19 or Influenza laboratory test |  |
| Comorbidities |  |  |
| Medical history prior to testing | POPULATION as defined **PLUS** sequential criteria of present diagnoses up to 20 years prior to COVID-19 or Influenza laboratory test |  |
| New-onset comorbidities after testing | Population as defined **AND** (exclusion of sequential criteria of present diagnosis up to 20 years before performed COVID-19 or Influenza laboratory test) **PLUS** sequential criteria of diagnosis up to 30 days after COVID-19 or Influenza laboratory test |  |
| **List of ICD-10 diagnosis codes used for identifying prior and new-onset comorbidities** |  |  |
| Neurological diagnosis, any (G00-99) |  |  |
| Cerebrovascular diagnosis (I60-69, G45 and G46) |  |  |
| Ischemic stroke (I63) |  |  |
| Psychiatric diagnosis, any (F00-99) |  |  |
| Ischemic heart disease (I20-25) |  |  |
| Heart failure (I50) |  |  |
| Diabetes, any (E10-14) |  |  |
| Chronic lower respiratory disease (J40-47) |  |  |
| Obesity (E66) |  |  |
| Anxiety (F40-48, neurotic, stress-related and somatoform disorders) |  |  |
| depression/affective disorder (F30-39, mood (affective) disorders) |  |  |
| schizophrenia (F20-29, Schizophrenia, schizotypal and delusional disorders) |  |  |
| Organic mental disorders (F00-09) |  |  |
| Delirium (F05) |  |  |
| Pulmonary embolism (I26) |  |  |
| Rheumatoid arthritis (M05-06, seropositive and other Rheumatoid arthritis) |  |  |
| Nephropathy (N17-19, Acute, chronic and unspecified renal failure) |  |  |
| Venous thrombosis (I80, all phlebitis and thrombophlebitis) |  |  |

## Validation of mortality data

To ensure verification of data, absolute mortality rates extracted from electronic health records (EPIC) were compared with official Danish statistics numbers:

**eTable 2.** Absolute mortality rates extracted from electronic health records (EPIC) and compared to official Danish numbers.

|  |  |  |
| --- | --- | --- |
| **Year** | **EPIC** | **Official Danish statistics data** |
| **2018 (Jan-Dec)** | 25,733 | 24,791 |
| **2019 (Jan-Dec)** | 25,992 | 24,377 |

EPIC-extracted general mortality data during years 2017-2019 corresponded well to Danish statistics mortality data (https://www.statistikbanken.dk/FOD507) covering the Capital (Greater Copenhagen, Bornholm) and Zealand Regions of Denmark. The discrepancy between the EPIC and official numbers is due to registration of e.g. tourists and patients from outside the Capital and Zealand Regions, treated for whatever reasons at a hospital in these 2 regions (which includes Rigshospitalet, Copenhagen University Hospital, a large tertiary referral center).

*Reference: Bliddal M, Broe A, Pottegård A, Olsen J, Langhoff-Roos J. The Danish Medical Birth Register. Eur. J. Epidemiol. 2018;33(1):27–36*.

**eTable 3.** Demographics and comorbidities in total populations (i.e. in- and outpatients combined)

|  | **COVID-19**<br>**(N = 7,920)** | **COVID-neg**<br>**(N=189,883)** | **OR (95%**<br>**CI)** | **p value** | **FLU**<br>**(N =**<br>**14,404)** | **OR (95%**<br>**CI)** | **p value** |
| --- | --- | --- | --- | --- | --- | --- | --- |
| **Age – years, no. (%)** |  |  |  |  |  |  |  |
| **Mean** | 51 | 50 |  |  | 58 |  |  |
| **18 - 40** | 2,492 (31.5) | 62,115 (32.7) | 0.94 (0.9-0.99) | 0.02 | 3,322 (23.1) | 1.53 (1.44-1.63) | < 0.001 |
| **40 - 60** | 2,936 (37.1) | 69,491 (36.6) | 1.02 (0.97-1.07) | 0.39 | 4,050 (28.1) | 1.50 (1.42-1.60) | < 0.001 |
| **60 - 80** | 1,679 (21.2) | 44,427 (2.4) | 0.88 (0.83-0.93) | < 0.001 | 4,637 (32.2) | 0.57 (0.53-0.60) | < 0.001 |
| **> 80** | 813 (10.3) | 13,850 (7.3) | 1.45 (1.35-1.57) | < 0.001 | 2,395 (16.6) | 0.57 (0.53-0.62) | < 0.001 |
| **Sex (%)** |  |  |  |  |  |  |  |
| **Women** | 4,666 (58.9) | 116,425 (61.3) | 0.90 (0.86-0.95) | < 0.001 | 8,116 (56.3) | 1.11 (1.05-1.17) | < 0.001 |
| **Prior medical diagnoses - no. (%)** |  |  |  |  |  |  |  |
| **Neurological, any** | 780 (9.8) | 19,072 (10.0) | 0.98 (0.91-1.06) | 0.57 | 1,499 (10.4) | 0.86 (0.79-0.95) | 0.002 |
| **Cerebrovascular, any** | 286 (3.6) | 6,517 (3.4) | 1.05 (0.93-1.19) | 0.39 | 610 (4.2) | 0.85 (0.73-0.98) | 0.023 |
| **Ischemic stroke** | 163 (2.1) | 4,041 (2.1) | 0.97 (0.82-1.13) | 0.67 | 295 (2.0) | 1.01 (0.83-1.22) | 0.05 |
| **Psychiatric, any** | 585 (7.4) | 18,845 (9.9) | 0.72 (0.66-0.79) | < 0.001 | 1,126 (7.8) | 0.94 (0.85-1.04) | 0.24 |
| **Ischemic heart disease** | 237 (3.0) | 6,350 (3.3) | 0.89 (0.78-1.02) | 0.09 | 702 (4.9) | 0.60 (0.52-0.70) | < 0.001 |
| **Heart failure** | 145 (1.8) | 3,442 (1.8) | 1.01 (0.85-1.19) | 0.91 | 490 (3.4) | 0.53 (0.44-0.64) | < 0.001 |
| **Diabetes** | 408 (5.2) | 8,194 (4.3) | 1.20 (1.09-1.33) | < 0.001 | 1,082 (7.5) | 0.67 (0.59-0.75) | < 0.001 |
| **Chronic lower respiratory disease** | 405 (5.1) | 11,877 (6.3) | 0.81 (0.73-0.89) | < 0.001 | 1,728 (12.0) | 0.40 (0.35-0.44) | < 0.001 |
| **Obesity** | 178 (2.2) | 5,225 (2.8) | 0.81 (0.7-0.95) | 0.007 | 308 (2.1) | 1.05 (0.87-1.27) | < 0.59 |

COVID-19: positive COVID-19 PCR test between March 1-June 1, 2020; FLU: positive influenza A/B PCR test between November 1, 2017 to March 1, 2020; COVID-neg: negative COVID-19 PCR test between March 1-June 1, 2020. Each patient was followed for a total of 30 days from positive test until end of follow-up or death. CI - confidence interval; no., number; OR, odds ratio. OR denotes comparisons of COVID-19 with study populations.

**eTable 4.** Demographics and comorbidities in in- and outpatient populations – OR and p-values compared to COVID-19 population as exposed group.

|  |  |  |  |  |  |  |  |  |
| --- | --- | --- | --- | --- | --- | --- | --- | --- |
|  | **Inpatients<sup>#</sup>** |  |  |  | **Outpatients<sup>#</sup>** |  |  |  |
|  | **COVID-neg**<br>**OR (95% CI)** | **COVID-neg**<br>**p-value** | **FLU**<br>**OR (95% CI)** | **FLU**<br>**p-value** | **COVID-neg**<br>**OR (95% CI)** | **COVID-neg**<br>**p-value** | **FLU**<br>**OR (95% CI)** | **FLU**<br>**p-value** |
| **Age – years** |  |  |  |  |  |  |  |  |
| **18 - 39** | 0.52 (0.45-0.61) | <0.001 | 0.83 (0.71-0.98) | 0.03 | 1.11 (1.05-1.17) | <0.001 | 1.17 (1.10-1.26) | <0.001 |
| **40 - 59** | 1.26 (1.13 - 1.41) | <0.001 | 1.42 (1.26-1.61) | <0.001 | 1.02 (0.97-1.07) | 0.45 | 1.16 (1.08-1.24) | <0.001 |
| **60 - 80** | 1.12 (1.01-1.24) | 0.03 | 0.94 (0.84-1.05) | 0.24 | 0.75 (0.71-0.81) | <0.001 | 0.62 (0.57-0.67) | <0.001 |
| **> 80** | 1.41 (1.26-1.58) | <0.001 | 0.87 (0.77-0.99) | 0.03 | 1.31 (1.18-1.45) | <0.001 | 1.12 (0.97-1.29) | 0.12 |
| **Sex** |  |  |  |  |  |  |  |  |
| **Women** | 0.68 (0.61-0.74) | <0.001 | 0.70 (0.63-0.78) | <0.001 | 1.00 (0.95-1.06) | 0.95 | 1.16 (1.08-1.24) | <0.001 |
| **Prior medical diagnoses - no. (%)** |  |  |  |  |  |  |  |  |
| **Neurological, any** | 1.05 (0.92-1.20) | 0.43 | 1.20 (1.04-1.39) | 0.01 | 0.86 (0.78-0.94) | 0.001 | 1.25 (1.09-1.42) | 0.001 |
| **Cerebrovascular, any** | 0.95 (0.79-1.13) | 0.54 | 1.29 (1.06-1.57) | 0.01 | 0.99 (0.83-1.17) | 0.92 | 1.34 (1.05-1.71) | 0.02 |
| **Ischemic stroke incl. TIA** | 0.84 (0.67-1.07) | 0.16 | 2.11 (1.60-2.77) | <0.001 | 0.93 (0.75-1.16) | 0.53 | 0.75 (0.57-0.99) | 0.04 |
| **Psychiatric, any** | 0.53 (0.45-0.61) | <0.001 | 1.08 (0.92-1.27) | 0.37 | 0.78 (0.70-0.86) | <0.001 | 1.47 (1.26-1.72) | <0.001 |
| **Ischemic heart disease** | 1.07 (0.90-1.27) | 0.46 | 1.18 (0.98-1.43) | 0.08 | 0.61 (0.49-0.75) | <0.001 | 0.66 (0.51-0.86) | 0.002 |
| **Heart failure** | 1.02 (0.83-1.26) | 0.84 | 1.03 (0.82-1.29) | 0.79 | 0.72 (0.53-0.97) | 0.03 | 0.76 (0.52-1.11) | 0.15 |
| **Diabetes** | 1.17 (1.00-1.36) | 0.04 | 1.05 (0.89-1.24) | 0.55 | 1.08 (0.94-1.24) | 0.28 | 0.94 (0.78-1.13) | 0.52 |
| **Chronic lower respiratory disease** | 0.86 (0.74-1.01) | 0.06 | 0.53 (0.45-0.62) | <0.001 | 0.70 (0.61-0.80) | <0.001 | 0.69 (0.58-0.82) | <0.001 |
| **Obesity** | 0.67 (0.49-0.90) | 0.007 | 1.20 (0.86-1.67) | 0.27 | 0.85 (0.71-1.01) | 0.06 | 1.08 (0.85-1.37) | 0.54 |
| COVID-neg: negative COVID-19 PCR test between March 1-June 1, 2020. FLU: positive influenza A/B PCR test between November 1, 2017 to March 1, 2020; Each patient was followed for a total of 30 days from positive test until end of follow-up or death. TIA: Transient ischemic attack. <sup>#</sup>at the time of testing |  |  |  |  |  |  |  |  |

**eTable 5.** Demographics and comorbidities of patients in COVID-19 population and subgroups.

|  | **COVID-19**<br>**(N = 7,920)** | **FLU-19**<br>**(N = 566)** | **FLU-19 neg**<br>**(N = 8,318)** | **FLU-18**<br>**(N = 3,838)** | **FLU-18 neg**<br>**(N = 11,153)** | **FLU-17**<br>**(N = 4,721)** | **FLU-17 neg**<br>**(N = 9,104)** |
| --- | --- | --- | --- | --- | --- | --- | --- |
| **Age – years, no. (%)** |  |  |  |  |  |  |  |
| **mean** | 51 | 55 | 60 | 55 | 59 | 59 | 61 |
| **18 - 40** | 2,492 (31.5) | 164 (29.0) | 1,570 (18.9) | 992 (25.8) | 2,388 (21.4) | 900 (19.1) | 1,834 (20.1) |
| **40 - 60** | 2,936 (37.1) | 145 (25.6) | 2,288 (27.5) | 1,215 (31.7) | 2,791 (25.0) | 1,367 (29.0) | 2,136 (23.5) |
| **60 - 80** | 1,679 (21.2) | 170 (30.0) | 2,995 (36.0) | 1,111 (28.9) | 3,998 (35.8) | 1,590 (33.7) | 3,281 (36.0) |
| **> 80** | 813 (10.3) | 87 (15.4) | 1,465 (17.6) | 520 (13.5) | 1,976 (17.7) | 864 (18.3) | 1,853 (20.4) |
| **Sex (%)** |  |  |  |  |  |  |  |
| **Women** | 4,666 (58.9) | 308 (54.4) | 4,500 (54.1) | 2,171 (56.6) | 6,220 (55.8) | 2,639 (55.9) | 5,087 (55.9) |
| **Prior medical history - no. (%)** |  |  |  |  |  |  |  |
| **Neurological, any** | 780 (9.8) | 84 (14.8) | 1,364 (16.4) | 453 (11.8) | 1,495 (13.4) | 427 (9.0) | 994 (10.9) |
| **Cerebrovascular** | 286 (3.6) | 43 (7.6) | 673 (8.1) | 158 (4.1) | 686 (6.2) | 175 (3.7) | 465 (5.1) |
| **Ischemic stroke** | 163 (2.1) | 26 (4.6) | 379 (4.6) | 68 (1.8) | 367 (3.3) | 84 (1.8) | 218 (2.4) |
| **Psychiatric, any** | 585 (7.4) | 48 (8.5) | 1,208 (14.5) | 368 (9.6) | 1,242 (11.1) | 307 (6.5) | 781 (8.6) |
| **Ischemic heart disease** | 237 (3.0) | 31 (5.5) | 686 (8.2) | 190 (5.0) | 756 (6.8) | 187 (4.0) | 545 (6.0) |
| **Heart failure** | 145 (1.8) | 23 (4.1) | 496 (6.0) | 135 (3.5) | 560 (5.0) | 146 (3.1) | 380 (4.2) |
| **Diabetes** | 408 (5.2) | 43 (7.6) | 864 (10.4) | 283 (7.4) | 1,020 (9.1) | 335 (7.1) | 761 (8.4) |
| **CLRD** | 405 (5.1) | 73 (12.9) | 1,500 (18.0) | 491 (12.8) | 1,865 (16.7) | 525 (11.1) | 1,414 (15.5) |
| **Obesity** | 178 (2.2) | 17 (3.0) | 204 (2.5) | 101 (2.6) | 274 (2.5) | 75 (1.6) | 179 (2.0) |
| **Admission – no. (%)** |  |  |  |  |  |  |  |
| **Inpatients** | 1,657 (20.9) | 317 (56.0) | 5,058 (60.8) | 1,842 (48.0) | 6,220 (55.8) | 2,376 (50.3) | 5,503 (60.4) |
| **Outpatients** | 6,263 (79.1) | 249 (44.0) | 3,260 (39.2) | 1,996 (52.0) | 4,933 (44.2) | 2,345 (49.7) | 3,601 (39.6) |

Basic demographics and comorbidities of study populations. COVID-19, Positive COVID-19 PCR test between March 1 to June 1, 2020; FLU-19, Positive influenza PCR test between March 1 to June 1, 2020; FLU-18, Positive influenza A/B PCR test between November 1, 2018 to March 1, 2019; FLU-17, Positive influenza A/B PCR test between November 1, 2017 to March 1, 2018; COVID-neg, Negative COVID-19 PCR test between March 1 to June 1, 2020. Patients with a positive or negative test were included in the given inclusion period. Each included patient was followed for a total of 30 days from positive test until end of follow-up or death. CLRD, Chronic lower respiratory disease; no., number; COVID, coronavirus disease

**eTable 6.** Relative risk of mortality, in- and outpatients in main populations, 30 days after test compared to COVID-19 negative individuals.

|  | Total inpatient (N) | Death inpatient (N) | Inpatient RR (95% CI) | Total outpatient (N) | Death outpatient (N) | Outpatient RR (95% CI) |
| --- | --- | --- | --- | --- | --- | --- |
| **COVID-19 positive** |  |  |  |  |  |  |
| **Overall** | 1,657 | 356 | 3.10 (2.80-3.42)<sup>†</sup> | 6,263 | 128 | 6.9 (5.69-8.38)<sup>†</sup> |
| **18-39 years** | 182 | <5 | N/A | 2,310 | <5 | N/A |
| **40-59 years** | 427 | 16 | 1.61 (0.97-2.67) | 2,509 | <5 | N/A |
| **60-80 years** | 636 | 150 | 2.65 (2.27-3.08)<sup>†</sup> | 1,043 | 33 | 6.52 (4.50-9.43)<sup>†</sup> |
| **> 80 years** | 412 | 190 | 2.73 (2.42-3.08)<sup>†</sup> | 401 | 92 | 6.27 (5.07-7.76)<sup>†</sup> |
| **COVID-19 negative** |  |  |  |  |  |  |
| **Overall** | 31,483 | 2,185 | 1 (ref) | 158,400 | 469 | 1 (ref) |
| **18-39 years** | 7,456 | 15 | 1 (ref) | 54,659 | <5 | 1 (ref) |
| **40-59 years** | 6,790 | 158 | 1 (ref) | 62,701 | 20 | 1 (ref) |
| **60-80 years** | 11,261 | 1,004 | 1 (ref) | 33,166 | 161 | 1 (ref) |
| **> 80 years** | 5,976 | 1,008 | 1 (ref) | 7,874 | 288 | 1 (ref) |
| **FLU positive** |  |  |  |  |  |  |
| **Overall** | 7,200 | 516 | 1.03 (0.94-1.13) | 7,204 | 27 | 1.27 (0.85-1.86) |
| **18-39 years** | 931 | <5 | N/A | 2,390 | <5 | N/A |
| **40-59 years** | 1,413 | 26 | 0.79 (0.52-1.19) | 2,638 | <5 | N/A |
| **60-80 years** | 2,876 | 214 | 0.83 (0.72-0.96)<sup>‡</sup> | 1,761 | <5 | N/A |
| **> 80 years** | 1,980 | 271 | 0.81 (0.72-0.92)<sup>‡</sup> | 415 | 20 | 1.32 (0.85-2.05) |
| **FLU negative** |  |  |  |  |  |  |
| **Overall** | 26,366 | 2,873 | 1.57 (1.49-1.66)<sup>†</sup> | 19,314 | 129 | 2.26 (1.86-2.74)<sup>†</sup> |
| **18-39 years** | 3,867 | 40 | 5.14 (2.84-9.30)<sup>†</sup> | 6,664 | <5 | N/A |
| **40-59 years** | 4,757 | 224 | 2.02 (1.66-2.47)<sup>†</sup> | 6,454 | 6 | 2.91 (1.17-7.25)<sup>‡</sup> |
| **60-80 years** | 10,939 | 1,307 | 1.34 (1.24-1.45)<sup>†</sup> | 4,963 | 51 | 2.12 (1.55-2.90)<sup>†</sup> |
| **> 80 years** | 6,803 | 1,302 | 1.13 (1.05-1.22)<sup>†</sup> | 1,233 | 71 | 1.57 (1.22-2.03)<sup>†</sup> |

Relative mortality rates divided in inpatients and outpatients. COVID-19 negative individuals were compared as the reference group with all other study populations. COVID-19, positive COVID-19 test between March 1-June 1, 2020; FLU positive, positive influenza A / B test between November 1, 2017-March 1, 2020; FLU negative, negative influenza A/B test between November 1, 2017-March 1, 2020; COVID-neg, negative COVID-19 test between March 1-June 1, 2020. Each included patient was followed for a total of 30 days from positive test until end of follow-up or death. ‡ p < 0.05; † p < 0.001; N/A, not applicable; ref, reference group

**eTable 7.** Mortality rates, 30 days after test in subgroups*.

|  | Total population | Inpatients |  | Outpatients |  |
| --- | --- | --- | --- | --- | --- |
|  | N | N (%) | Death (%) | N (%) | Death (%) |
| **COVID-19 positive** | 7,920 | 1,657 (20.9) | 356 (21.5) | 6,263 (79.1) | 128 (2.0) |
| **COVID-19 negative** | 189,883 | 31,483 (16.6) | 2,185 (1.4) | 158,400 (83.4) | 469 (0.3) |
| **FLU positive** | 14,404 | 7,200 (50.0) | 516 (7.2) | 7,204 (50.0) | 27 (0.4) |
| **FLU negative** | 45,680 | 26,366 (57.7) | 2,873 (11) | 19,314 (42.3) | 129 (0.7) |
| **FLU-19 positive** | 566 | 317 (56.0) | 26 (8.2) | 249 (44.0) | ≤5 |
| **FLU-19 negative** | 8,318 | 5,058 (60.8) | 578 (11.4) | 3,260 (39.2) | 74 (2.3) |
| **FLU-18 positive** | 3,838 | 1,842 (48.0) | 121 (6.6) | 1,996 (52.0) | 9 (0.5) |
| **FLU-18 negative** | 11,153 | 6,220 (55.8) | 649 (10.4) | 4,933 (44.2) | 41 (0.8) |
| **FLU-17 positive** | 4,721 | 2,376 (50.3) | 174 (7.3) | 2,345 (49.7) | ≤5 |
| **FLU-17 negative** | 9,104 | 5,503 (60.4) | 597 (10.8) | 3,601 (39.6) | 21 (0.6) |

Absolute mortality rates of study populations divided in inpatients and outpatients. COVID-19, positive COVID-19 test between March 1-June 1, 2020; FLU-18, positive influenza A/B test between November 1, 2018-March 1, 2019; FLU-17, positive influenza A/B test between November 1, 2017-March 1, 2018; COVID-neg, negative COVID-19 test between March 1-June 1, 2020. Each included patient was followed for 30 days from positive test until end of follow-up or death.\* Data presented as ≤5 is due to the General Data Protection Regulation law restricting display of results with 5 or fewer patients, thus no statistical analysis was performed.

**eTable 8.** Relative risk of mortality, in- and outpatients in subgroups, 30 days after test compared to COVID-19 positive individuals as exposed group.

| Pairwise comparisons | Inpatient, RR (95% CI) | Outpatient, RR (95% CI) |
| --- | --- | --- |
| COVID-19-positive | 3.10 (2.80-3.42)† | 6.90 (5.69-8.38)† |
| COVID-19-negative | 1 (ref) | 1 (ref) |
| COVID-19-positive | 2.62 (1.79-3.83)† | N/A |
| FLU-19-positive | 1 (ref) | N/A |
| COVID-19-positive | 2.62 (1.79-3.83)† | 0.90 (0.68-1.20) |
| FLU-19-negative | 1 (ref) | 1 (ref) |
| COVID-19-positive | 3.27 (2.69-3.98)† | 4.53 (2.31-8.89)† |
| FLU-18-positive | 1 (ref) | 1 (ref) |
| COVID-19-positive | 2.06 (1.83-2.32)† | 2.46 (1.73-3.49)† |
| FLU-18-negative |  |  |
| COVID-19-positive | 2.93 (2.47-3.48)† | N/A |
| FLU-17-positive | 1 (ref) | N/A |
| COVID-19-positive | 1.98 (1.76-2.23)† | 3.50 (2.21-5.55)† |
| FLU-17-negative | 1 (ref) | 1 (ref) |

Relative mortality rates divided in inpatients and outpatients. COVID-19 positive individuals were compared as the exposed group with all other populations. COVID-19, positive COVID-19 test between March 1-June 1, 2020; FLU-18, positive influenza A/B test between November 1, 2018-March 1, 2019; FLU-17, positive influenza A/B test between November 1, 2017-March 1, 2018; COVID-neg, negative COVID-19 test between March 1-June 1, 2020. Each included patient was followed for 30 days from positive test until end of follow-up or death. N/A, not applicable. † p < 0.001

**eTable 9.** Relative risk of mortality, in- and outpatients, 30 days after test compared to COVID-19 negative individuals as reference group.

| Study populations | Inpatient, RR (95% CI) | Outpatient, RR (95% CI) |
| --- | --- | --- |
| COVID-19 negative | 1 (ref) | 1 (ref) |
| COVID-19 positive | 3.10 (2.80-3.42)† | 6.9 (5.69-8.38)† |
| FLU-19 positive | 1.18 (0.82-1.71) | N/A |
| FLU-19 negative | 1.65 (1.51-1.80)† | 7.67 (6.01-9.77)† |
| FLU-18 positive | 0.95 (0.79-1.13) | 1.52 (0.79-2.94) |
| FLU-18 negative | 1.50 (1.38-1.63)† | 2.81 (2.04-3.76)† |
| FLU-17 positive | 1.06 (0.91-1.22) | N/A |
| FLU-17 negative | 1.56 (1.43-1.70)† | 1.96 (1.27-3.05)‡ |

Relative mortality rates divided in inpatients and outpatients. COVID-19 negative individuals were compared as the reference group with all other study populations. COVID-19, positive COVID-19 test between March 1-June 1, 2020; FLU-18, positive influenza A / B test between November 1, 2018-March 1, 2019; FLU-17, positive influenza A/B test between November 1, 2017-March 1, 2018; COVID-neg, negative COVID-19 test between March 1-June 1, 2020. Each included patient was followed for a total of 30 days from positive test until end of follow-up or death. ‡ p < 0.01; † p < 0.001; N/A, not applicable

**eTable 10.** New-onset diagnosis, 30-days after COVID-19 test*.

| **eTable 10. New-onset diagnosis, 30-days after COVID-19 test\*** |  |  |  |  |
| --- | --- | --- | --- | --- |
|  | **COVID-19**<br>**(N = 7,920)** | **COVID-neg**<br>**(N=189,883)** | **RR (95% CI)** | **p value** |
| New diagnosis after 30 days - no. (%)# |  |  |  |  |
| **Cerebrovascular disorders (any)** | 22 (0.3) | 681 (0.4) | 0.78 (0.51-1.86) | 0,2409 |
| **Ischemic stroke** | 15 (0.2) | 423 (0.2) | 0.85 (0.51-1.42) | 0,5356 |
| **Ischemic heart disease** | 11 (0.1) | 497 (0.3) | 0.53 (0.29-0.96) | 0.0364 |
| **Pulmonary embolism** | 23 (0.3) | 224 (0.1) | 2.47 (1.60-3.78) | <0.0001 |
| **Venous thrombosis** | 9 (0.1) | 126 (0.1) | 1.71 (0.87-3.36) | 0.1194 |
| **Nephropathy** | 43 (0.6) | 235 (0.1) | 4.41 (3.19-6.10) | <0.0001 |
| **Diabetes** | 45 (0.6) | 335 (0.2) | 3.25 (2.38-4.43) | <0.0001 |
| **Rheumatoid arthritis** | ≤5 | 18 (0.01) | N/A | N/A |
| **Neurological disorders (excluding vascular)** | 16 (0.2) | 799 (0.5) | 0.48 (0.29-0.79) | 0,0035 |
| **Psychiatric disorders (any)** | 24 (0.3) | 593 (0.3) | 0.94 (0.63-1.42) | 0,7806 |
| **Organic mental disorders** | 11 (0.1) | 152 (0.1) | 1.75 (0.95-3.23) | 0,0719 |
| **Delirium** | ≤5 | 91 (0.05) | N/A | N/A |
| **Neurotic and anxiety disorders** | ≤5 | 211 (0.1) | N/A | N/A |
| **Affective disorders** | ≤5 | 145 (0.1) | N/A | N/A |
| **Schizophrenia and delusional disorders** | ≤5 | 84 (0.04) | N/A | N/A |

New-onset diagnoses in all COVID-19 and influenza A/B positive and negative individuals, 30 days after testing. RR denotes relative risk (RR) of new-onset diagnoses comparisons of COVID-19 positive with COVID-19 negative individuals. COVID-19, positive COVID-19 test between March 1-June 1, 2020; COVID-neg, negative COVID-19 test between March 1-June 1, 2020. Patients were followed for 30 days from positive test until end of follow-up or death. \* Data presented as ≤5 is due to the General Data Protection Regulation law restricting display of results with 5 or fewer patients, thus no statistical analysis was performed. # Percentage is calculated as new-onset diagnosis divided by sum of population excluding individuals with prior history of the investigated diagnosis.

**eTable 11.**
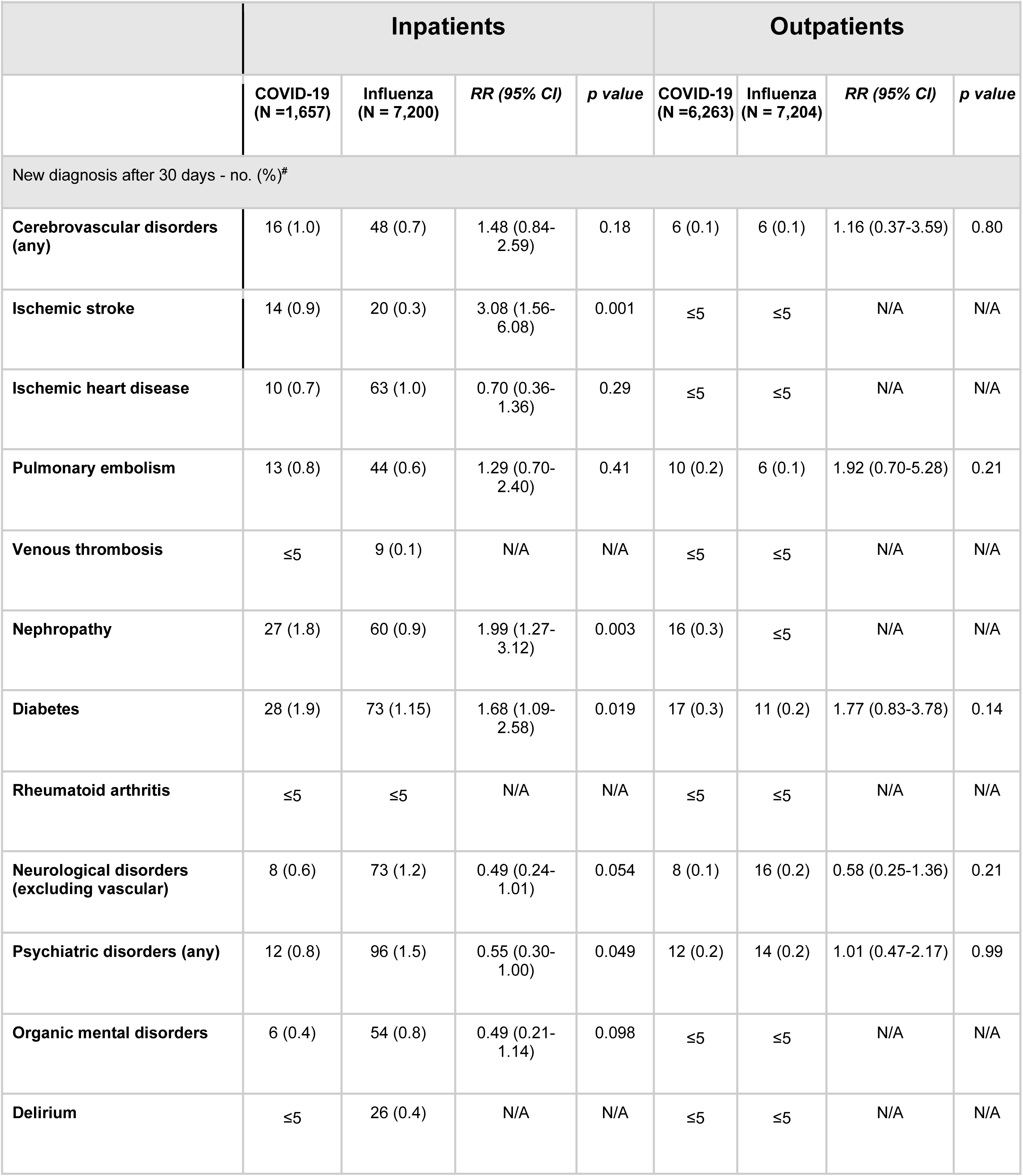
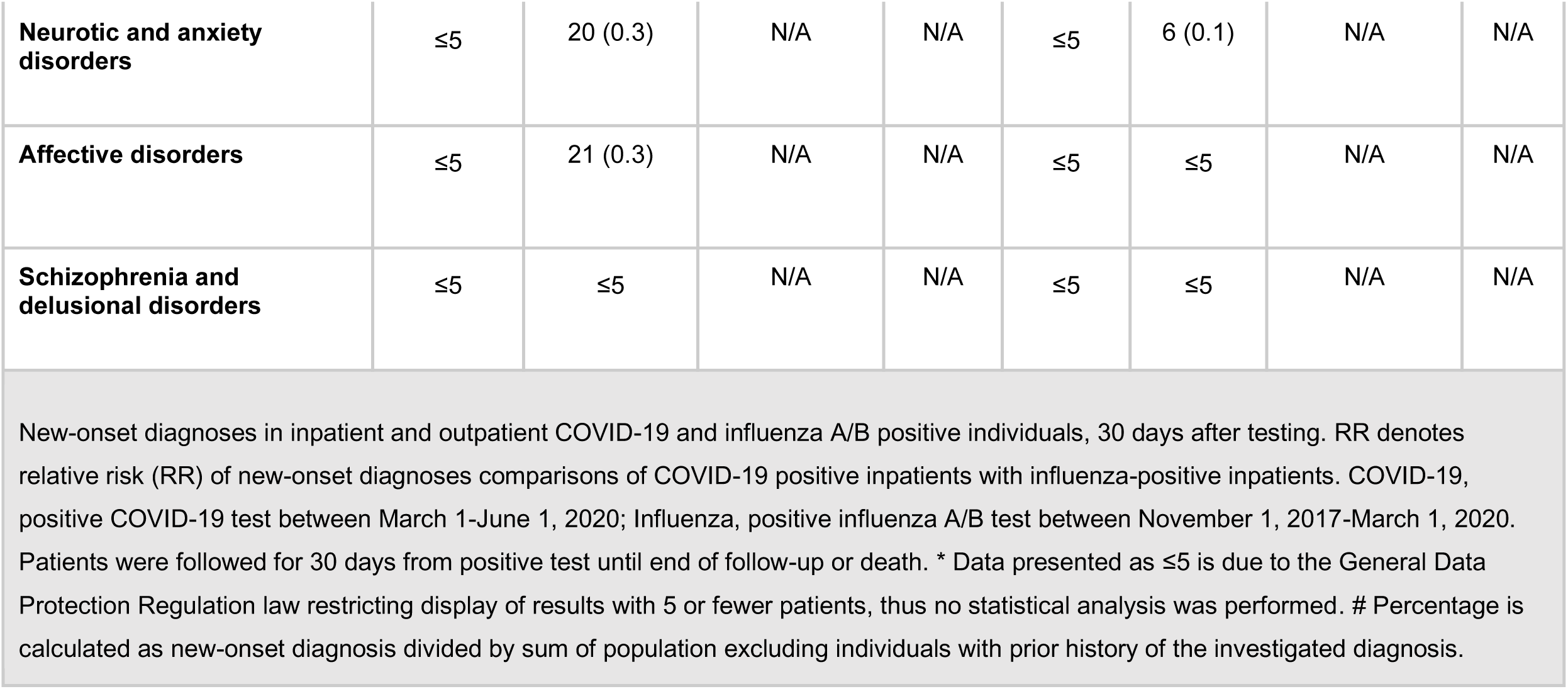
New-onset diagnosis in inpatients and outpatients, 30-days after positive COVID-19 or influenza A/B test*.

